# Interleukin 21 is a marker of Human African Trypanosomiasis Infection and a contributor to pathology in mice

**DOI:** 10.1101/2024.05.07.24306988

**Authors:** Paul Capewell, Hamidou Ilboudo, Anneli Cooper, Windingoudi Justin Kaboré, Harry Noyes, Kerry O’ Neill, Mamadou Camara, Vincent Jamonneau, Annette MacLeod, Bruno Bucheton, TrypanoGEN+ research group of the H3Africa consortium

## Abstract

**Background:** Human African trypanosomiasis (HAT) is an important disease of sub-Saharan Africa that is approaching elimination in many regions. However, the disease has previously returned from similarly low case numbers in the past, making it important to identify issues that hinder elimination efforts. One important factor is likely to be the recent characterization of individuals with latent HAT infections that are able to tolerate HAT with few symptoms and to control blood parasitaemia to levels that are undetectable by microscopy. Although animal trypanotolerance has been examined in detail, it is unclear how the latent phenotype is maintained in humans.

**Methods:** To identify immune components involved in latent HAT, we used targeted RNASeq to examine the expression of 495 immune-related transcripts in blood collected from 287 individuals at active disease foci in Guinea. These samples included latent infections, HAT clinical cases, and uninfected controls. The *in vivo* effects of IL21 functional blockade was investigated using a murine model of trypanosomiasis.

**Results:** Differential expression analysis revealed transcripts involved in T cell activation and B cell development that associated with trypanosome infection, including *PD1*, *CD70*, and *CD80*. In particular, *IL21* was found to be elevated in infected individuals, although it was significantly higher in clinical cases relative to latent infections. This pattern was replicated at the protein level when patient sera were examined by ELISA. Reducing IL21 pathway activity in mice infected with *Trypanosoma brucei* led to increased survivorship and reduced parasitaemia in the model animals.

**Conclusion:** Our data show that IL21 is a potential biomarker of Human African Trypanosomiasis and is a cause rather than a consequence of symptoms severity. Further investigation of IL21 will contribute to understanding the factors involved in developing latent HAT, improving control efforts to identify and predict such infections. In the future, the factors identified in this study may also serve as intervention targets to control the symptoms of trypanosomiasis.

## Introduction

Human African trypanosomiasis (HAT) is an important vector-borne disease of sub-Saharan Africa primarily caused by the parasite *Trypanosoma brucei gambiense (T. b. gambiense)* [1]. The related veterinary disease animal trypanosomiasis (AT) is also a significant economic burden involving a complex of trypanosome species, including *T. congolense*, *T. vivax*, and *T. b. brucei* [2]. HAT has been targeted for elimination as a public health problem by 2020 by the World Health Organization (WHO) and reported cases have fallen below 1,000 in recent years [1,3]. The next objective on the HAT control road map is to reach the interruption of transmission by 2030. However, several issues have recently been identified that could hinder these efforts. These include the description of individuals able to tolerate the disease with few symptoms which has overturned of the long-held belief that HAT is invariably fatal. These individuals also display low blood parasitemias and despite positive responses to the serological screening tests, they are generally left untreated by the control programs as the microscopic confirmation of trypanosomes is required in order to initiate treatment [4–8]. These latent infections are initially identified by an elevated response to the card agglutination test for trypanosomiasis (CATT) and then further examined using an immune trypanolysis test targeting the LiTAT1.3 Variable Surface Glycoprotein known to be highly specific for *T.b. gambiense* in humans [9]. These individuals, termed SERO TL+, are definitively characterised as harbouring a latent infection only if they maintain serological positivity for at least two years without developing symptoms while remaining negative to parasitological tests performed on biological fluids [5,6]. During a two-year longitudinal study led in Guinea,13% of SERO TL+ individuals developed symptoms with detectable parasites (termed SERO TL+/HAT), 48% remained latent [5] and 39% became negative to the CATT in a probable process of selfcure [5]. The latent period can be extremely protracted, with one case persisting for 29 years before an unrelated immunosuppressive treatment led to rapid onset of HAT [10]. It has also recently been revealed that African trypanosomes are not exclusively parasites of the blood and can exists in tissues of the host. Most notably, they can colonise the skin where they could act as a significant but overlooked anatomical reservoir [11–13]. Modelling suggests it is likely that these latent infections carry infective parasites and are able to maintain HAT in disease foci without animal reservoirs despite intensive control efforts [14]. This is supported by xenodiagnosis experiments examining asymptomatic people [15] and animals [16] demonstrating that the tsetse vector can be infected by latent hosts and more recently by the detection of trypanosomes in skin biopsies sampled in Guinea from both confirmed HAT patients but also serologically positive individuals that displayed ortherwise negative parasitology in blood or lymph [17]. The genotypes of parasites from SERO TL+ individuals are indistinguishable from those infecting clinical cases [7] and genomic analyses of *T. b. gambiense* isolates from across Africa show that the parasite is clonal and highly homogenous [18]. Together, these data suggest that the host is the primary mediator of the differences in symptom severity that can lead to latent infections and SERO TL+ individuals.

Investigations of the *APOL1* G1 and G2 polymporphisms within the TrypanoGEN network evidenced a genetic association between the G2 polymorphism and resistance to *T. b. rhodesiense* infection [19,20] as well as an increased frequency of G1 carriage in individuals with latent *T. b. gambiense* infections [20][21]. Several immunological studies targeted to specific immune mediators have also revealed differences in the immune response between SERO TL+ and HAT patients. IL8 was elevated in SERO TL+ and in these individuals was predictive of the CATT becoming negative during the follow-up. In addition immunosuppressive molecules such as IL10 and the maternal human leukocyte antigen protein HLA-G were found more elevated in HAT patients and SERO TL+ individuals who subsequently developed the disease [22,23]. However, no causative links between immune factors and latent infections have been demonstrated and the design of earlier studies to decipher the mechanisms of infection resistance/susceptibility in human have had to largely rely on data from studies of trypanotolerant animals. Trypanotolerance has been extensively studied in mice and cattle [24,25], although the majority of such studies have used the animal trypanosome species *T. congolenese*, *T. vivax*, and *T. b. brucei* rather than *T. b. gambiense*. The term trypanotolerance is also somewhat misleading as these animals do become infected with trypanosomes, develop adverse effects and mount an immune response. Trypanotolerance should rather be understood as reduced pathology and disease in the presence of infection. Despite the different parasite species used, studies of trypanotolerant animals suggest some factors that may affect disease presentation in humans [26]. These include the importance of a strong IFNg-mediated response early in infection to control the initial peak of parasitaemia, followed by reduction in the IFNg-mediated inflammatory environment that leads to a long-term and less virulent infection. Key to mediating this environmental shift are the cytokines IL10 and IL4. A robust B cell and humoral response is also required to control the parasite, particularly during the initial peak of parasitaemia [27].

To more fully understand the immune components involved in HAT symptom severity and the development of human latent infections, we compared the peripheral blood transcriptomes of *T. b. gambiense* HAT clinical cases, people with latent infections and uninfected controls using a targeted immune transcript analysis. This revealed transcripts involved in T cell activation and B cell development that differed between clinical and latent cases. More specifically, expression of the cytokine *IL21* at both the transcript and protein level associated with symptom severity. Subsequent *in vivo* experiments using functional blockade confirmed that IL21 is involved in determining both symptom severity and parasitaemia during trypanosome infection. This cytokine therefore serves as both a biomarker of trypanosome infection and also offers a new intervention target to limit symptoms of the disease in humans and animals.

## Methods

### Study Design

Participants were identified during medical surveys organised by the Guinean National Control Programme (NCP) of HAT from three HAT foci in Guinea (Dubreka, Boffa, and Forecariah) according to the WHO and NCP policies [5,28]. Five mL samples of blood were collected in heparinised tubes from individuals who tested positive for infection using the card agglutination trypanosomiasis test (CATT). A 2-fold dilution series of patient blood was tested to estimate CATT titre. Individuals with titres 1/4 or greater were submitted for microscopic examination of lymph node aspirates and 350 µl of buffy coat were examined for the presence of trypanosomes using mini-anion exchange centrifugation test (mAECT). If trypanosomes were detected, lumbar puncture was performed, and disease stage determined by identifying trypanosomes and by white blood cell (WBC) counts. Confirmed HAT cases were classified as stage 1 (0–5 WBC/µL), early stage 2 (6–20 WBC/µL; or ≤20 WBC with trypanosomes in CSF), or late stage 2 (>20 WBC/µL) and treated according to NCP guidelines [28]. For each study individual, 100 µL of plasma was sampled and 2 mL of blood were taken on PAXgene^®^ Blood RNA tubes (PreAnalytiX). All samples were frozen in the field at -20°C in car freezer and were then stored at -80 °C until use. The plasma was used to perform the immune trypanolysis test to detect Litat 1.3 and Litat 1.5 variable surface antigens specific for *T. b. gambiense* [9]. The PAXgene^®^ blood RNA tubes were used to extract RNA with PAXgene^®^ Blood RNA kit (PreAnalytiX). CATT and trypanolysis positive subjects without visible parasites were defined as SERO TL+. SERO TL+ individuals were followed-up for at least two years (with an average of three visits) and of these, 11 went on to develop HAT and were defined as SERO TL+/HAT. Those that remained CATT and trypanolysis positive for at least two years were defined as having latent infections.

### Targeted RNASeq

A Qiagen Human Inflammation & Immunity Transcriptome Kit (RHS-005Z; Qiagen) was used to obtain transcript counts from a panel of 495 genes involved in the innate and acquired immune responses (Supplementary File 1). cDNA libraries were prepared from 56 uninfected controls, 168 HAT cases (135 pre- and 33 post-treatment), 36 SERO TL+ samples with latent infections and 11 individuals who were intitially CATT and TL positive but parasitologically negative but became parasitologically positive on follow up (SERO/HAT). The resulting Illumina libraries were sequenced on an Illumina NextSeq 500 and filtered and mapped using Qiagen GeneGlobe software (Qiagen). Normalization and pairwise differential expression analysis for each transcript were performed by Qiagen using DESeq2 for R [29] and normalised read counts are provided as Supplementary Data File 4.

### IL21 ELISA

A human IL21 ELISA kit (ab119542; Abcam) was used to measure IL21 titres in sera from eight samples randomly selected from each group (Control, HAT, SERO TL+, and SERO/HAT) following the manufacturer’s instructions. There were three replicates for each sample.

### Anti-IL21 intervention

To assess the effects of reducing IL21 pathway function *in vivo*, 8-week-old female BALB/c mice were housed in a specific pathogen-free environment in accordance with local and Home Office regulations. Two experiments were performed using four mice treated with 100 μg recombinant anti-IL21 neutralizing antibody every other day from day -1 (FFA21; Thermo Fisher Scientific). These concentrations were selected based on previous *in vivo* studies examining the role of IL21 during viral infection [30]. In addition, four mice per experiment were injected with 100 μg isotype antibody on the same days to serve as controls (Rat IgG2a κ; Thermo Fisher Scientific). All mice were inoculated intraperitoneally with 10^4^ STIB247 *T. b. brucei* parasites on day 0 and parasitaemia was assayed on each subsequent day using phase microscopy [31].

## Results

### Targeted RNASeq

In order to identify potential differentiators between infected individuals (both SERO TL+ and HAT) and controls, in addition to clinical cases and latent individuals, a cost-effective targeted RNASeq approach was used to examine 495 immune-related genes in whole blood RNA purified from 271 individuals from active disease foci in Guinea. The targeted genes encompass various canonical B cell and T cell pathways (Supplementary File 1). The sample library included uninfected controls (n = 56); HAT cases pre-treatment (n = 135); post-treatment HAT cases (n = 33); latent infections (SERO TL+ individuals who had remained serology positive and parasitologically negative for two years) (n = 36) and SERO/HAT (individuals who were seropositive but parasitiologically negative at first screening but became parasitologically positive at a later screening) (n = 11). Normalised read counts for all genes and participants are available in Supplementary data file 4.The expression profiles of these groups were then compared pairwise to reveal differentially expressed transcripts (Supplementary 3; Figure 1). In a comparison of HAT cases versus controls, 10 transcripts were found to be differentially regulated with an FDR *p_*adj < 0.001 (Figure 1A). Expression of the high affinity IgE receptor *FCER1A* was lower in HAT cases, but the expression levels of *IL21*, *CD1C*, *TLR10*, *IL12B*, *PD1*, *CD70*, *CD80*, *CAV1*, and *MET* were all higher. No significant differences were found between HAT cases post-treatment and controls, although a comparison of HAT cases pre- and post-treatment revealed that the expression levels of *IL21*, *CD1C*, *IL10*, *IL12B*, *CAV1*, and *MET* were all significantly higher in the pre-treatment group (*p*_adj < 0.001; Figure 1B). A comparison of individuals with latent infections versus controls found only two transcripts that differed significantly in expression (*IL21* and *IL12B*) with *p*_adj < 0.001 (Figure 1C). Both of these transcripts were also significant in the HAT versus control comparison. Finally, a comparison of individuals with latent infections and HAT clinical cases found four transcripts that showed significant differences in expression (*CD1C*, *IL21*, *CAV1*, and *MET*) with *p*_adj < 0.001 (Figure 1D). Again, these transcripts were significant in the HAT clinical cases versus control comparison. It was noted that *IL21* was significant across multiple comparisons and elevated in both HAT and latent infection groups. However, expression was significantly higher in the clinical HAT group relative to the latent infection cohort, suggesting it may be involved in the development of HAT symptoms. To establish if the pattern observed in *IL21* transcript expression correlated with the protein level, an anti-IL21 ELISA was performed using sera from a subset of samples from each group (n = 8 for each group). This confirmed that the pattern of IL21 expression at the protein level was the same as the transcript level, being elevated in infected individuals but much higher in clinical HAT (Figure 2). Finally, to assess if IL21 levels may be predictive for SERO TL+ individuals that go on to develop HAT, titres from the 11 SERO/HAT individuals were also examined. These samples were taken from patients that were serologically positive without symptoms but went on to develop disease with detectable parasitaemia during the two-year follow-up period. This differs from the SERO TL+ group that maintained their latent phenotype with undetectable parasitaemia. IL21 titres in the SERO/HAT group that progressed to active disease were not found to be significantly different from SERO TL+ individuals that maintained their latent phenotype for the duration of the study, suggesting that elevated IL21 is associated with infection rather than acting as a predictive marker of risk of developing active disease. Nevertheless, these data reveal that *IL21* transcript and IL21 protein are elevated during trypanosome infection and are higher in clinical HAT cases relative to latent SERO TL+ infections.

**Figure 1.**
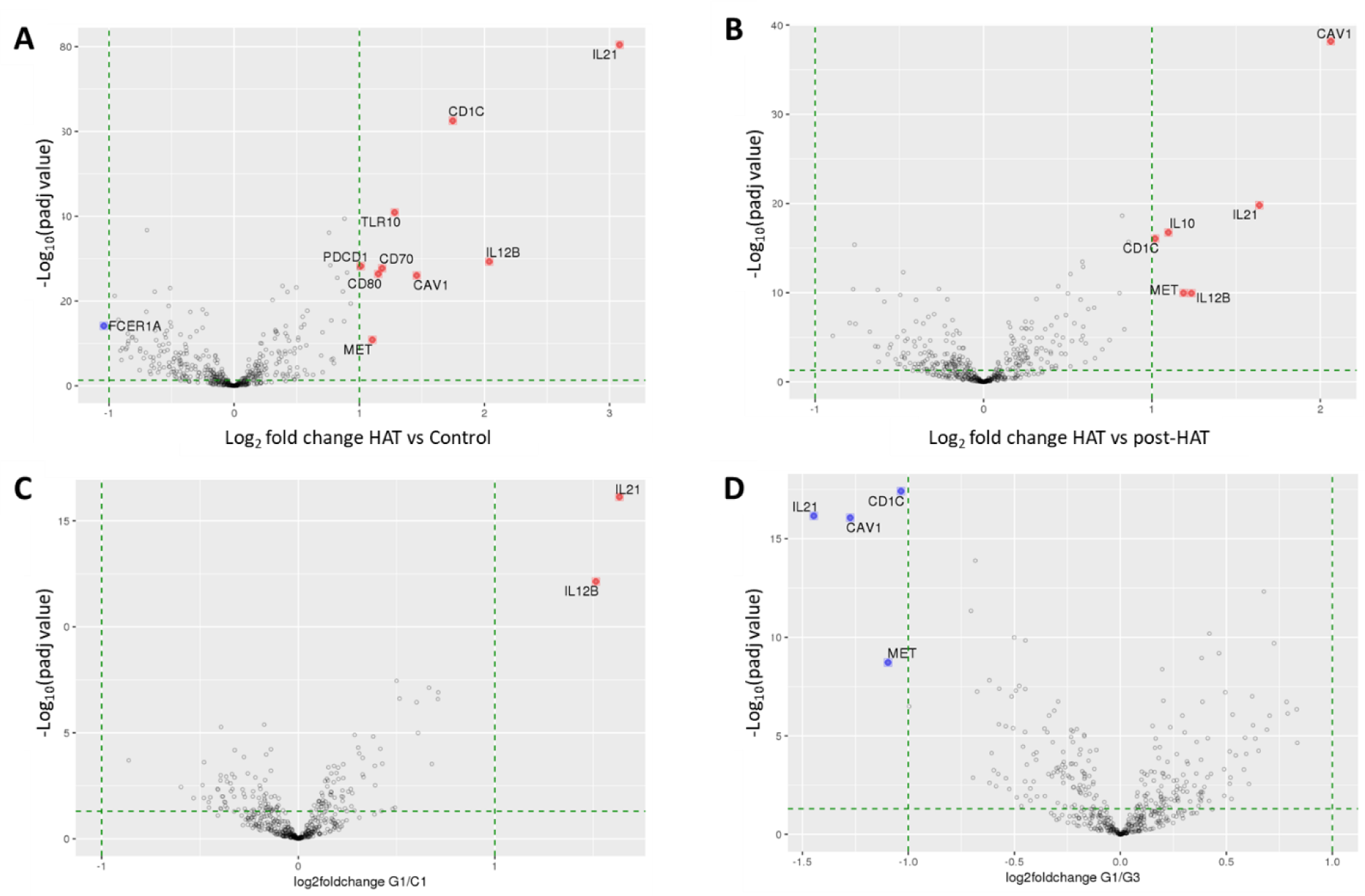
Genes with differential expression between latent infections, HAT, and Control groups. Volcano plots demonstrating the relationships between -log_10_(*p*_adj values) and the log_2_ fold change values obtained from DESeq2. Genes showing differential expression with a *p_*adj value above 0.05 and log_2_ fold change ≤ -1 or ≥ 1 are coloured red (up-regulated) or blue (down-regulated). The panels show significant genes identified in comparisons between (A) clinical HAT cases versus controls, (B) clinical HAT cases versus HAT post-treatment, (C) individuals with latent infections versus controls, and (D) individuals with latent infections versus HAT cases.

**Figure 2.**
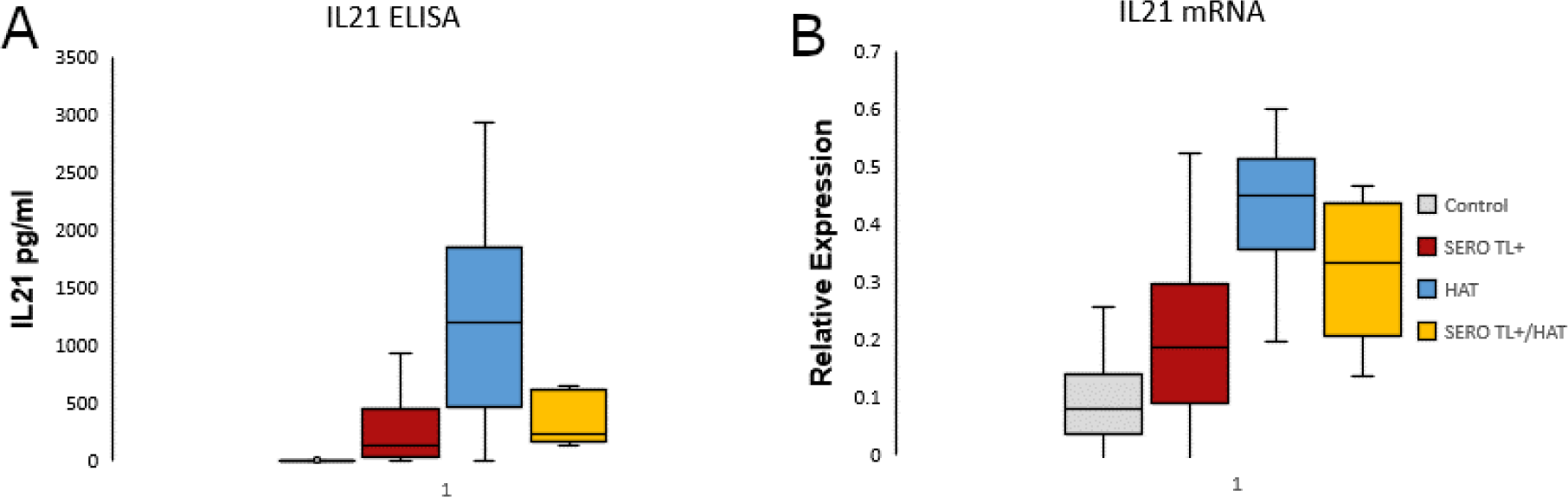
Serum IL21 Titres and mRNA expression show that both protein and mRNA levels increase after infection. **(A)** IL21 titres were assessed by ELISA for a random subset of control, latent infections (SERO TL+), individuals with HAT and those who have developed active HAT after first presenting with just positive serology (SERO/HAT). Sera were from the targeted RNASeq study (n = 8 per group). There was a significant difference between both SERO TL+ and SERO/HAT individuals and HAT patients (T-Test, p < 0.01) but no difference between SERO TL+ and SERO/HAT individuals (T test, p = 0.90). **(B)** *IL21* mRNA from the targeted RNAseq data representing the same groups.

### IL21 Functional Intervention

To establish whether IL21 was a cause or consequence of HAT severity, the IL21 pathway was inhibited using a neutralising anti-IL21 antibody in a HAT mouse model across two trials (parasitaemia data are presented in Supplementary File 2). During the first trial, one animal in the control group treated with isotype control succumbed to trypanosomiasis at day eight and the remaining controls were euthanized due to severe illness. All of the mice in the experimental group treated with the neutralising antibody were healthy and presented with no symptoms, although they were also euthanized due to the terms of the license under which the work was performed. During the second trial, the experiment was again curtailed at day eight due to severe symptom and paralysis in the control group. As in the previous experiment all of the mice treated with anti-IL21 antibody were visibly healthy. This suggests that the IL21 pathway is involved in increased symptom severity during trypanosome infection rather than being simply a consequence of symptom development. However, the early end of these experiment means further work is required to fully understand the mechanisms involved. In addition, when the parasitaemia curves from the two independent experiments were aligned to the first peak of parasitaemia, a significant interaction effect was observed between treatment and day (ANOVA, F = 3.02, p < 0.01). An interaction effect represents the combined effects of factors on the dependent measure (parasitaemia), with the impact of one factor depending on the level of another factor. In this case, the effect of the neutralising anti-IL21 antibody on parasitaemia depended on days post-infection (Figure 3). When days were examined individually, there was a significant difference observed in the magnitude of the first peak of parasitaemia between treated and control groups (T Test, p = 0.03). These data show that administration of the anti-IL21 antibody affected parasite number in addition to affecting symptoms, demonstrating that IL21 has a significant impact on parasitaemia and symptom severity during African trypanosome infections. In particular, reducing IL21 pathway function associates with better outcomes and lower parasitaemia.

**Figure 3.**
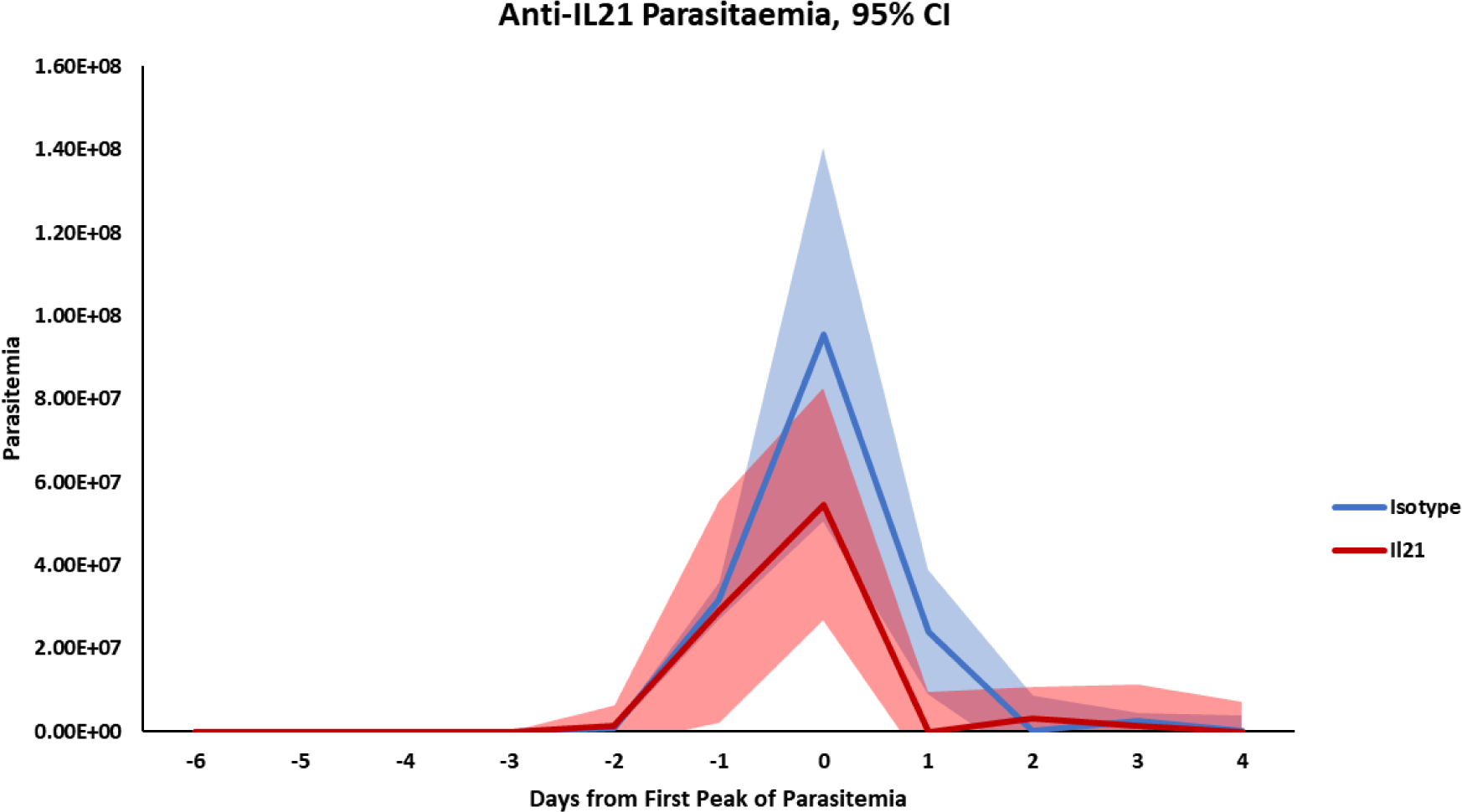
The effect of IL21 functional blockade on parasitaemia in a HAT model. Eight mice were infected with *T. b. brucei* STIB247 across two different experiments. Mice were administered with a blocking anti-IL21 chimera antibody (red line) or an isotype control (blue line) on alternate days beginning the day before inoculation. Parasitaemia was estimated daily via microscopy and the resulting curves were aligned to the first peak (day 0) to account for individual variation. Blocking IL21 consistently reduced parasitiaemia. Mean parasitaemia for treatment and isotype control are shown with 95% confidence intervals. A significant interaction effect was observed between treatment and days post-infection (ANOVA, p < 0.01) and there was a significant difference observed in size of the first parasitaemia peak (T Test, p = 0.03).

## Discussion

As HAT approaches elimination, it becomes increasingly important to identify and understand factors that may impact the control of this deadly parasite, particularly latent infections, and the role of the skin as an anatomical reservoir [3,32]. Our study used RNASeq on a panel of 496 genes involved in the innate and acquired immune responses to identify immune components involved in the latent phenotype in 260 individuals from HAT active foci in Guinea. When comparing HAT cases and controls, we identified 10 transcripts that associate with clinical disease, some of which have never previously been identified as being involved in HAT. These included transcripts for *IL21*, *IL12B*, *IL10, CD1C*, *TLR10*, *PD1*, *CD70*, *CD80*, *CAV1*, and *MET*. Interleukin-12 subunit beta (IL12B) is a component of IL12, a cytokine involved in a wide range of immune responses during infectious challenge [33,34]. However, the subunit is also a component of IL23, and our data is unable to assign this increase in *IL12B* expression to IL12, IL23, or both. However, IL23 is an important mediator of Th17-mediated and GM-CSF responses involved in controlling extracellular parasites [35]. Many of the protective roles assigned to IL12 are now also being reassessed in light of the description of IL23 [36]. Further work is therefore required to establish whether IL23 expression is important during African trypanosome infection and to establish the role of Th17 responses. Transcripts involved in T cell activation and exhaustion were also identified, including *PD1* and *CD70* (both immune checkpoint markers), and *CD80* (a receptor for the co-stimulatory protein CD28). The involvement of T cell activation and co-stimulation imply that increased T cell activation is occurring in infected individuals, likely in response to the presence of a pathogen. However, it has also been suggested that African trypanosomes possess a lymphocyte triggering factor (TLTF) that drives polyclonal (non-specific) T activation [37,38], leading to higher expression of proteins associating with immune exhaustion and suppression. PD1, highlighted in our study, is one such suppressive protein that acts via its ligands (PDL1 and PDL2) to down-regulate the immune system and promote self-tolerance [39]. Although PD1 has not previously been associated with trypanosome infection, the pathway has been implicated in the response and control of several important parasites, including *Toxoplasma*, *Leishmania*, and *Plasmodium* [40–42]. Both PD1 and IL10 were elevated in HAT cases and they co-operate to suppress CD8+ Tcells [43]. Severe immune suppression is a classically described feature of African trypanosome infection in humans and livestock [44][45] and we hypothesize that this may be due to increased activation of IL10 and the PD1 pathway in infected hosts. Further investigation is required to establish if this is the case and whether intervention can alleviate immune suppression and improve symptoms, as shown for *Plasmodium* [41] and *Leishmania donovani* [42].

Very little is known about Toll-like receptor 10 (TLR10) as mice do not possess the protein, limiting investigation [46]. However, unlike all other TLRs, it is immune suppressive and may therefore be involved in the systemic immune suppression seen during trypanosome infections [47]. Our data indicate that African trypanosomes may serve as a powerful model system to elucidate some of the mechanisms of these understudied immune proteins. Finally, our comparison between HAT cases and controls revealed that the high affinity IgE receptor *FCER1A* showed lower expression in infected patients. Previously it has been shown in trypanosome-infected rats that there is a reduced anaphylaxis due to parasite-mediated disruption of IgE-specific pathways [48]. Our data suggest a similar effect may occur in humans and that lowered expression of *FCER1A* may contribute to a decrease in IgE responses. It is unclear why African trypanosomes would target these anaphylactic pathways, but it may be related to controlling the immune response in the skin. Previous data has shown that there is almost no inflammatory reaction to trypanosomes present in the skin of infected mice, despite high parasite burdens [12]. Finally, our data show that after treatment, the expression of these transcripts, including immune suppressive transcripts such as *PD1,* return to levels equivalent to controls. This suggests that the perturbation to a patient’s immune system is not permanent. A similar effect has been observed in the immune systems of HAT patients that have been previously examined [49].

Across each of the targeted RNASeq comparisons, *IL21* has emerged as a recurring factor that was highlighted as elevated in both HAT cases and latent infections versus controls. However, *IL21* transcript levels were lower in latent cases when compared to HAT patients. This suggests that *IL21* is elevated during trypanosome infection but is significantly higher in clinical cases. This pattern was confirmed at the protein level by ELISA. Our subsequent *in vivo* intervention using a mouse model of HAT has confirmed that reducing the activity of the IL21 pathway through functional blockade leads to higher survival rates and reduced parasitaemia in infected animals. This demonstrates that *IL21* expression influences the severity of symptoms rather than elevated expression simply being a consequence of African trypanosome infection. In contrast blockade of IL21 or IL21R in mice infected with *Plasmodium chabaudi* lead to sustained high parasitaemia and a failure to resolve the chronic stage of the infection [50]. IL21 is a pleiotropic cytokine with a wide range of interactions across immune and non-immune cell types [51]. It is primarily made by activated T cells, particularly T follicular helper (Tfh cells), and one of its primary functions is to promote B cell development and maintain humoral responses [52]. Patients with defective IL21 signalling are severely immune compromised, suffering from deficiencies in plasma cell and memory B cell generation, as well as immunoglobulin class switching [53–55]. They also suffer from repeated secondary infections due to increased susceptibility to infection [54]. However, IL21 has been described as a “double-edged sword” due to a complicated role in immunity, either supporting and suppressing B cell development depending on serum levels and interactions with other cytokines present [51][56]. In particular, IL21 has a potent immunosuppressive effect at high serum titres, primarily through Tfh driving cell cycle arrest and apoptosis in B cells [57–59]. Interestingly, destruction of the B cell compartment is a distinctive component of African trypanosome infection in mice [60], although this effect may be less pronounced during more chronic *T. b. gambiense* infections [61]. Nevertheless, our data suggest that disruption of the B cell population in mice may be due to elevated IL21 levels and further experimentation is required to confirm this hypothesis. Although we did not observe a decrease in B cell numbers during our *in vivo* experiments (data not shown), these data were collected on day eight and B cell destruction was not noticeable until after day 10 in previous studies [60].

Based on our data, we hypothesize that a perturbation in T cell activation and B cell development contributes to immune suppression, higher parasitaemia, and the development of symptoms in clinical HAT cases. IL21 appears intimately involved in these affects and our data suggest that latent infections with reduced symptoms are in some part due to the control of this cytokine. As IL21 is primarily produced by activated T cells, reducing activation would lead to less activity in the IL21 pathway, maintaining a robust B cell and humoral response. In a previous study that examined trypanosome-mediated polyclonal T cell activation in susceptible and resistance mice, more resistant strains differed in their response to the polyclonal activator TLTF by increasing expression of cytokines associated with antibody synthesis and class switching [62]. While this supports our hypothesis, much more investigation is required to establish which cell populations are expressing IL21 and whether blockade of activation checkpoint proteins (such as PD1) can alleviate symptoms in an animal model of HAT. In *Plasmodium* infections, polyclonal T cell activation caused by the parasite leads to elevated expression of PD1 on Tfh cells, again leading to impaired B cell and humoral responses. When PD1-mediated suppression is alleviated via blocking antibodies (alongside blockade of another checkpoint marker LAG3), Tfh function and B cell responses are restored, resulting in less severe symptoms [41]. It is possible that a similar effect is occurring during HAT, with individuals with latent infections displaying less activation and exhaustion in the Tfh population, maintaining B cell and humoral responses, leading to a long term control of parasitaeimia but not parasite elimination. This would suggest that IL21 and PD1 are potential intervention targets that may play a role in controlling symptoms by maintaining the strength of the host immune system, particularly in livestock where the elimination of trypanosomiasis is unlikely if not impossible.

In addition, of its potential functional role in mediating *T. b. gambiense* infection outcomes, IL21 also appears as an interesting biomarker of trypanosome infection in human. With the upcoming availability of new oral drugs, in particular Acoziborol given as a single dose, the treatment of individuals testing positive to the actual serological tests (CATT or RDTs) is likely to become a reality in the coming years if the drug is proved to be safe enough [63,64]. A major challenge of this largely simplified test and treat strategy reside in the fact that all the actual antibody detection tests for HAT lack specificity and in low endemic prevalence contexts display poor positive predictive values [65]. Detection of IL21 in a simple test format in those serological suspects is likely to enhance the identification of individuals with active infections thus reducing considerably the number of “unnecessary” treatments. The fact that the plasma levels of IL21 were shown to be significantly higher in both latent and HAT patients as compared to controls is a proof of concept that it is a reachable goal. Larger multi-centric studies integrating other co-infections as potential confounding factors will be required to further validate IL21 as a biomarker *T. b. gambiense* infection in humans.

## Conclusion

Our study is an example of successfully applying targeted transcriptomics to identify genes associating with latent carriage of HAT, highlighting IL21 as a potential biomarker of infection as well as a key mediator of symptom severity and a likely contributor to the development of a latent phenotype. Importantly, we have used an animal model to confirm the relevance of IL21 during African trypanosome infections. Further investigation of IL21, and its role in disease, will improve our understanding of the factors involved in developing clinical HAT or latent infections, but may also contribute to the development of more productive and trypanosome tolerant livestock breeds.

## Data Availability

All data produced in the present study are available upon reasonable request to the authors and the TrypanoGEN network (H3Africa)

## Declarations

### Ethics approval and consent to participate

All investigations on humans were conducted in accordance with the Declaration of Helsinki. Participants were identified through healthcare providers, community engagement and active surveillance campaigns led by the national contral program, Ministry of Health Guinea. Written informed consents for sample collection, analysis and publication of anonymised data was obtained from all participants by trained local healthcare workers. Subjects or their legal guardian gave consent as a signature or a thumbprint after receiving standardized information in French or their local langage as preferred. Ethical approvals for the study was obtained from within the TrypanoGEN Project following H3Africa Consortium guidelines for informed consent [66] and from Comité Consultatif de Déontologie et d’éthique (CCDE) at the Institut de recherché pour le Développent (IRD; 10/06/2013). Research procedures were also approved by the University of Glasgow MVLS Ethics Committee for Non-Clinical Research Involving Human Subjects (Reference no. 200120043).

All animal experiments were approved by the University of Glasgow Ethical Review Committee and performed in accordance with the UK Home Office guidelines, UK Animals (Scientific Procedures) Act, 1986 and EU directive 2010/63/EU. All experiments were conducted under SAPO regulations and UK Home Office project licence number PC8C3B25C to Dr. Jean Rodgers.

### Competing interests

The authors declare that they have no competing interests

### Funding

PC, AC, AML were funded by a Wellcome Senior Fellowship to AML (209511/Z/17/Z). BB was funded by IRD. WJK, HN and HI, were supported through the Human Hereditary and Health in Africa (H3Africa) [H3A/18/004]. The second phase of the Wellcome component of H3Africais being implemented by the African Academy of Sciences (AAS) and the NEPAD Agency’s Alliance for Accelerating Excellence in Science in Africa (AESA) in partnership with Wellcome.

### Authors’ contributions

PC conducted in vitro and vivo experiments drafted manuscript; AC conducted in vitro and in vivo experiments; WJK Sample Collection and RNA prep revised manuscript; HI Sample Collection and RNA prep; HN Revised manuscript; KO’N In vivo experiments; MC, VJ Managed and participated in sample collection; AM and BB Devised experiment and revised manuscript.

## Acknowledgements

The authors would like to acknowledge the study participants who donated their specimens, the personnel involved in the community engagement and coordinating sample collection and processing, the national sleeping sickness control programmes of the participating countries. The views expressed in this publication are those of the author(s) and not necessarily those of AAS, the NEPAD Agency nor Wellcome.

## Notes

### Competing Interest Statement

The authors have declared no competing interest.

### Author Declarations

All investigations on humans were conducted in accordance with the Declaration of Helsinki. Participants were identified through healthcare providers, community engagement and active surveillance campaigns led by the national contral program, Ministry of Health Guinea. Written informed consents for sample collection, analysis and publication of anonymised data was obtained from all participants by trained local healthcare workers. Subjects or their legal guardian gave consent as a signature or a thumbprint after receiving standardized information in French or their local langage as preferred. Ethical approvals for the study was obtained from within the TrypanoGEN Project following H3Africa Consortium guidelines for informed consent and from Comite Consultatif de Deontologie et ethique (CCDE) at the Institut de recherche pour le Developpent (IRD; 10/06/2013). Research procedures were also approved by the University of Glasgow MVLS Ethics Committee for Non-Clinical Research Involving Human Subjects (Reference no. 200120043). All animal experiments were approved by the University of Glasgow Ethical Review Committee and performed in accordance with the UK Home Office guidelines, UK Animals (Scientific Procedures) Act, 1986 and EU directive 2010/63/EU. All experiments were conducted under SAPO regulations and UK Home Office project licence number PC8C3B25C to Dr. Jean Rodgers.

## References

1. WHO. Trypanosomiasis, human African (sleeping sickness). 2020 [cited 20 Apr 2022] p. WHO Fact Sheet, No. 259. Available: https://www.who.int/news-room/fact-sheets/detail/trypanosomiasis-human-african-(sleeping-sickness)

2. Kristjanson PM, Swallow BM, Rowlands GJ, Kruska RL, De Leeuw PN. Measuring the costs of African animal trypanosomosis, the potential benefits of control and returns to research. Agricultural Systems. 1999;59: 79–98. doi:10.1016/S0308-521X(98)00086-9

3. Akazue PI, Ebiloma GU, Ajibola O, Isaac C, Onyekwelu K, Ezeh CO, et al. Sustainable Elimination (Zero Cases) of Sleeping Sickness: How Far Are We from Achieving This Goal? Pathogens 2019, Vol 8, Page 135. 2019;8: 135. doi:10.3390/PATHOGENS8030135

4. Ilboudo H, Bras-Goncalves R, Camara M, Flori L, Camara O, Sakande H, et al. Unravelling human trypanotolerance: IL8 is associated with infection control whereas IL10 and TNFalpha are associated with subsequent disease development. PLoS Pathog. 2014;10: e1004469. doi:10.1371/journal.ppat.1004469

5. Ilboudo H, Jamonneau V, Camara M, Camara O, Dama E, Léno M, et al. Diversity of response to Trypanosoma brucei gambiense infections in the Forecariah mangrove focus (Guinea): Perspectives for a better control of sleeping sickness. Microbes and Infection. 2011. doi:10.1016/j.micinf.2011.05.007

6. Jamonneau V, Ilboudo H, Kaboré J, Kaba D, Koffi M, Solano P, et al. Untreated Human Infections by Trypanosoma brucei gambiense are Not 100% Fatal. PLoS Negl Trop Dis. 2012. doi:10.1371/journal.pntd.0001691

7. Kabore J, Koffi M, Bucheton B, MacLeod A, Duffy C, Ilboudo H, et al. First evidence that parasite infecting apparent aparasitemic serological suspects in human African trypanosomiasis are Trypanosoma brucei gambiense and are similar to those found in patients. Infect Genet Evol. 2011;11: 1250–1255. doi:10.1016/j.meegid.2011.04.014

8. Koffi M, Solano P, Denizot M, Courtin D, Garcia A, Lejon V, et al. Aparasitemic serological suspects in Trypanosoma brucei gambiense human African trypanosomiasis: a potential human reservoir of parasites? Acta Trop. 2006;98: 183–188. doi:10.1016/j.actatropica.2006.04.001

9. Jamonneau V, Bucheton B, Kaboré J, Ilboudo H, Camara O, Courtin F, et al. Revisiting the immune trypanolysis test to optimise epidemiological surveillance and control of sleeping sickness in West Africa. PLoS Neglected Tropical Diseases. 2010. doi:10.1371/journal.pntd.0000917

10. Sudarshi D, Lawrence S, Pickrell WO, Eligar V, Walters R, Quaderi S, et al. Human African Trypanosomiasis Presenting at Least 29 Years after Infection—What Can This Teach Us about the Pathogenesis and Control of This Neglected Tropical Disease? PLOS Neglected Tropical Diseases. 2014;8: e3349. doi:10.1371/JOURNAL.PNTD.0003349

11. Caljon G, Van Reet N, De Trez C, Vermeersch M, Pérez-Morga D, Van Den Abbeele J. The Dermis as a Delivery Site of Trypanosoma brucei for Tsetse Flies. PLOS Pathogens. 2016;12: e1005744. doi:10.1371/JOURNAL.PPAT.1005744

12. Capewell P, Cren-Travaille C, Marchesi F, Johnston P, Clucas C, Benson RA, et al. The skin is a significant but overlooked anatomical reservoir for vector-borne African trypanosomes. eLife. 2016;5. doi:10.7554/eLife.17716

13. Trindade S, Rijo-Ferreira F, Carvalho T, Pinto-Neves D, Guegan F, Aresta-Branco F, et al. Trypanosoma brucei Parasites Occupy and Functionally Adapt to the Adipose Tissue in Mice. Cell host & microbe. 2016;19: 837–848. doi:10.1016/J.CHOM.2016.05.002

14. Capewell P, Atkins K, Weir W, Jamonneau V, Camara M, Clucas C, et al. Resolving the apparent transmission paradox of African sleeping sickness. PLOS Biology. 2019;17: e3000105. doi:10.1371/JOURNAL.PBIO.3000105

15. Frezil JL. Application of xenodiagnosis for the demonstration of T. gambiense trypanosomiasis in immunologically suspect individuals. In: Bulletin de la Société de Pathologie Exotique [Internet]. 1971 [cited 20 Apr 2022] p. 64: 871-878. Available: https://www.cabdirect.org/cabdirect/abstract/19732900264

16. Wombou Toukam CM, Solano P, Bengaly Z, Jamonneau V, Bucheton B. Experimental evaluation of xenodiagnosis to detect trypanosomes at low parasitaemia levels in infected hosts. Parasite (Paris, France). 2011;18: 295–302. doi:10.1051/PARASITE/2011184295

17. Camara M, Soumah AM, Ilboudo H, Travaillé C, Clucas C, Cooper A, et al. Extravascular Dermal Trypanosomes in Suspected and Confirmed Cases of gambiense Human African Trypanosomiasis. Clinical Infectious Diseases. 2021;73: 12–20. doi:10.1093/cid/ciaa897

18. Weir W, Capewell P, Foth B, Clucas C, Pountain A, Steketee P, et al. Population genomics reveals the origin and asexual evolution of human infective trypanosomes. eLife. 2016;5. doi:10.7554/ELIFE.11473

19. Kamoto K, Noyes H, Nambala P, Senga E, Musaya J, Kumwenda B, et al. Association of APOL1 renal disease risk alleles with Trypanosoma brucei rhodesiense infection outcomes in the northern part of Malawi. PLOS Neglected Tropical Diseases. 2019;13: e0007603. doi:10.1371/journal.pntd.0007603

20. Cooper A, Ilboudo H, Alibu VP, Ravel S, Enyaru J, Weir W, et al. APOL1 renal risk variants have contrasting resistance and susceptibility associations with African trypanosomiasis. Tishkoff S, editor. eLife. 2017;6: e25461. doi:10.7554/eLife.25461

21. Kaboré JW, Ilboudo H, Noyes H, Camara O, Kaboré J, Camara M, et al. Candidate gene polymorphisms study between human African trypanosomiasis clinical phenotypes in Guinea. PLOS Neglected Tropical Diseases. 2017;11: e0005833. doi:10.1371/journal.pntd.0005833

22. Courtin D, Milet J, Sabbagh A, Massaro JD, Castelli EC, Jamonneau V, et al. HLA-G 3’ UTR-2 haplotype is associated with Human African trypanosomiasis susceptibility. Infection, Genetics and Evolution. 2013. doi:10.1016/j.meegid.2013.03.004

23. Gineau L, Courtin D, Camara M, Ilboudo H, Jamonneau V, Dias FC, et al. Human Leukocyte Antigen-G: A Promising Prognostic Marker of Disease Progression to Improve the Control of Human African Trypanosomiasis. Clinical Infectious Diseases. 2016. doi:10.1093/cid/ciw505

24. Courtin D, Berthier D, Thevenon S, Dayo GK, Garcia A, Bucheton B. Host genetics in African trypanosomiasis. Infect Genet Evol. 2008;8: 229–238. doi:10.1016/j.meegid.2008.02.007

25. Kemp SJ, Darvasi A, Soller M, Teale AJ. Genetic control of resistance to trypanosomiasis. Veterinary immunology and immunopathology. 1996;54: 239–243. doi:10.1016/S0165-2427(96)05692-9

26. Mansfield JM, Paulnock DM. Regulation of innate and acquired immunity in African trypanosomiasis. Parasite immunology. 2005;27: 361–371. doi:10.1111/J.1365-3024.2005.00791.X

27. Magez S, Schwegmann A, Atkinson R, Claes F, Drennan M, De Baetselier P, et al. The role of B-cells and IgM antibodies in parasitemia, anemia, and VSG switching in Trypanosoma brucei-infected mice. PLoS pathogens. 2008;4. doi:10.1371/JOURNAL.PPAT.1000122

28. Camara M, Kaba D, KagbaDouno M, Sanon JR, Ouendeno FF, Solano P. [Human African trypanosomiasis in the mangrove forest in Guinea: epidemiological and clinical features in two adjacent outbreak areas]. Med Trop (Mars). 2005;65: 155–161.

29. Love MI, Huber W, Anders S. Moderated estimation of fold change and dispersion for RNA-seq data with DESeq2. Genome Biology. 2014;15: 550. doi:10.1186/s13059-014-0550-8

30. Shen Z, Yang H, Yang S, Wang W, Cui X, Zhou X, et al. Hepatitis B virus persistence in mice reveals IL-21 and IL-33 as regulators of viral clearance. Nature communications. 2017;8. doi:10.1038/S41467-017-02304-7

31. Herbert WJ, Lumsden WHR. Trypanosoma brucei: a rapid “matching” method for estimating the host’s parasitemia. Experimental parasitology. 1976;40: 427–431. doi:10.1016/0014-4894(76)90110-7

32. Berthier D, Brenière SF, Bras-Gonçalves R, Lemesre JL, Jamonneau V, Solano P, et al. Tolerance to Trypanosomatids: A Threat, or a Key for Disease Elimination? Trends in parasitology. 2016;32: 157–168. doi:10.1016/J.PT.2015.11.001

33. Dorman SE, Holland SM. Interferon-gamma and interleukin-12 pathway defects and human disease. Cytokine & growth factor reviews. 2000;11: 321–333. doi:10.1016/S1359-6101(00)00010-1

34. Locksley RM. Interleukin 12 in host defense against microbial pathogens. Proceedings of the National Academy of Sciences of the United States of America. 1993;90: 5879–5880. doi:10.1073/PNAS.90.13.5879

35. Tang C, Chen S, Qian H, Huang W. Interleukin-23: as a drug target for autoimmune inflammatory diseases. Immunology. 2012;135: 112–124. doi:10.1111/J.1365-2567.2011.03522.X

36. Cua DJ, Sherlock J, Chen Y, Murphy CA, Joyce B, Seymour B, et al. Interleukin-23 rather than interleukin-12 is the critical cytokine for autoimmune inflammation of the brain. Nature. 2003;421: 744–748. doi:10.1038/NATURE01355

37. Abdulla MH, Bakhiet M, Lejon V, Andersson J, McKerrow J, Al-Obeed O, et al. TLTF in cerebrospinal fluid for detection and staging of T. b. gambiense infection. PloS one. 2013;8. doi:10.1371/JOURNAL.PONE.0079281

38. BAKHIET M, OLSSON T, EDLUND C, HÖJEBERG B, HOLMBERG K, LORENTZHN J, et al. A Trypanosoma brucei brucei-derived factor that triggers CD8+ lymphocytes to interferon-gamma secretion: purification, characterization and protective effects in vivo by treatment with a monoclonal antibody against the factor. Scandinavian journal of immunology. 1993;37: 165–178. doi:10.1111/J.1365-3083.1993.TB01753.X

39. Francisco LM, Sage PT, Sharpe AH. The PD-1 pathway in tolerance and autoimmunity. Immunological reviews. 2010;236: 219–242. doi:10.1111/J.1600-065X.2010.00923.X

40. Bhadra R, Gigley JP, Weiss LM, Khan IA. Control of Toxoplasma reactivation by rescue of dysfunctional CD8+ T-cell response via PD-1-PDL-1 blockade. Proceedings of the National Academy of Sciences of the United States of America. 2011;108: 9196–9201. doi:10.1073/PNAS.1015298108

41. Butler NS, Moebius J, Pewe LL, Traore B, Doumbo OK, Tygrett LT, et al. Therapeutic blockade of PD-L1 and LAG-3 rapidly clears established blood-stage Plasmodium infection. Nature immunology. 2011;13: 188–195. doi:10.1038/NI.2180

42. Habib S, El Andaloussi A, Elmasry K, Handoussa A, Azab M, Elsawey A, et al. PDL-1 Blockade Prevents T Cell Exhaustion, Inhibits Autophagy, and Promotes Clearance of Leishmania donovani. Infection and immunity. 2018;86. doi:10.1128/IAI.00019-18

43. Sun Z, Fourcade J, Pagliano O, Chauvin J-M, Sander C, Kirkwood JM, et al. IL10 and PD-1 Cooperate to Limit the Activity of Tumor-Specific CD8+ T Cells. Cancer Research. 2015;75: 1635– 1644. doi:10.1158/0008-5472.CAN-14-3016

44. Roelants GE, Pinder M. Immunobiology of African trypanosomiasis. Contemporary topics in immunobiology. 1984;12: 225–274. doi:10.1007/978-1-4684-4571-8_7

45. Marañón C. Whole genome transcriptome reveals metabolic and immune susceptibility factors for Trypanosoma congolense infection in West-African livestock. PCI Infections. 2023; 100008. doi:10.24072/pci.infections.100008

46. Chuang TH, Ulevitch RJ. Identification of hTLR10: a novel human Toll-like receptor preferentially expressed in immune cells. Biochimica et biophysica acta. 2001;1518: 157–161. doi:10.1016/S0167-4781(00)00289-X

47. Hess NJ, Jiang S, Li X, Guan Y, Tapping RI. TLR10 Is a B Cell Intrinsic Suppressor of Adaptive Immune Responses. Journal of immunology (Baltimore, Md : 1950). 2017;198: 699–707. doi:10.4049/JIMMUNOL.1601335

48. Gould SS, Castro GA. Suppression by Trypanosoma brucei of anaphylaxis-mediated ion transport in the small intestine of rats. Immunology. 1994/03/01 ed. 1994;81: 468.

49. Lejon V, Mumba Ngoyi D, Kestens L, Boel L, Barbé B, Kande Betu V, et al. Gambiense Human African Trypanosomiasis and Immunological Memory: Effect on Phenotypic Lymphocyte Profiles and Humoral Immunity. PLoS Pathogens. 2014;10. doi:10.1371/journal.ppat.1003947

50. Pérez-Mazliah D, Ng DHL, Rosário APF do, McLaughlin S, Mastelic-Gavillet B, Sodenkamp J, et al. Disruption of IL-21 Signaling Affects T Cell-B Cell Interactions and Abrogates Protective Humoral Immunity to Malaria. PLOS Pathogens. 2015;11: e1004715. doi:10.1371/journal.ppat.1004715

51. Leonard WJ, Wan CK. IL-21 Signaling in Immunity. F1000Research. 2016;5. doi:10.12688/F1000RESEARCH.7634.1

52. de Leur K, Dor FJMF, Dieterich M, van der Laan LJW, Hendriks RW, Baan CC. IL-21 receptor antagonist inhibits differentiation of B cells toward plasmablasts upon alloantigen stimulation. Frontiers in Immunology. 2017;8: 306. doi:10.3389/FIMMU.2017.00306/FULL

53. Erman B, Bilic I, Hirschmugl T, Salzer E, Çagdas D, Esenboga S, et al. Combined immunodeficiency with CD4 lymphopenia and sclerosing cholangitis caused by a novel loss-of-function mutation affecting IL21R. Haematologica. 2015;100: e216. doi:10.3324/HAEMATOL.2014.120980

54. Kotlarz D, Ziȩtara N, Uzel G, Weidemann T, Braun CJ, Diestelhorst J, et al. Loss-of-function mutations in the IL-21 receptor gene cause a primary immunodeficiency syndrome. The Journal of Experimental Medicine. 2013;210: 433. doi:10.1084/JEM.20111229

55. Stepensky P, Keller B, Abuzaitoun O, Shaag A, Yaacov B, Unger S, et al. Extending the clinical and immunological phenotype of human interleukin-21 receptor deficiency. Haematologica. 2015;100: e72. doi:10.3324/HAEMATOL.2014.112508

56. Spolski R, Leonard WJ. Interleukin-21: a double-edged sword with therapeutic potential. Nature reviews Drug discovery. 2014;13: 379–395. doi:10.1038/NRD4296

57. Jin H, Carrio R, Yu A, Malek TR. Distinct activation signals determine whether IL-21 induces B cell costimulation, growth arrest, or Bim-dependent apoptosis. Journal of immunology (Baltimore, Md : 1950). 2004;173: 657–665. doi:10.4049/JIMMUNOL.173.1.657

58. Mehta DS, Wurster AL, Whitters MJ, Young DA, Collins M, Grusby MJ. IL-21 induces the apoptosis of resting and activated primary B cells. Journal of immunology (Baltimore, Md : 1950). 2003;170: 4111–4118. doi:10.4049/JIMMUNOL.170.8.4111

59. Ozaki K, Spolski R, Ettinger R, Kim H-P, Wang G, Qi C-F, et al. Regulation of B cell differentiation and plasma cell generation by IL-21, a novel inducer of Blimp-1 and Bcl-6. Journal of immunology (Baltimore, Md : 1950). 2004;173: 5361–5371. doi:10.4049/JIMMUNOL.173.9.5361

60. Bockstal V, Guirnalda P, Caljon G, Goenka R, Telfer JC, Frenkel D, et al. T. brucei infection reduces B lymphopoiesis in bone marrow and truncates compensatory splenic lymphopoiesis through transitional B-cell apoptosis. PLoS pathogens. 2011;7. doi:10.1371/JOURNAL.PPAT.1002089

61. Cnops J, Kauffmann F, De Trez C, Baltz T, Keirsse J, Radwanska M, et al. Maintenance of B cells during chronic murine Trypanosoma brucei gambiense infection. Parasite immunology. 2016;38: 642–647. doi:10.1111/PIM.12344

62. M B, T O, A L, B H, P VDM, K K. Induction of interferon-gamma, transforming growth factor-beta, and interleukin-4 in mouse strains with different susceptibilities to Trypanosoma brucei brucei. Journal of interferon & cytokine research : the official journal of the International Society for Interferon and Cytokine Research. 1996;16: 427–433. doi:10.1089/JIR.1996.16.427

63. Franco JR, Cecchi G, Paone M, Diarra A, Grout L, Ebeja AK, et al. The elimination of human African trypanosomiasis: Achievements in relation to WHO road map targets for 2020. PLoS neglected tropical diseases. 2022;16. doi:10.1371/JOURNAL.PNTD.0010047

64. Dickie EA, Giordani F, Gould MK, Mäser P, Burri C, Mottram JC, et al. New Drugs for Human African Trypanosomiasis: A Twenty First Century Success Story. Tropical medicine and infectious disease. 2020;5. doi:10.3390/TROPICALMED5010029

65. Koné M, Kaba D, Kaboré J, Thomas LF, Falzon LC, Koffi M, et al. Passive surveillance of human African trypanosomiasis in Côte d’Ivoire: Understanding prevalence, clinical symptoms and signs, and diagnostic test characteristics. PLoS neglected tropical diseases. 2021;15. doi:10.1371/JOURNAL.PNTD.0009656

66. “H3Africa Working Group on Ethics and Regulatory Issues for the Human Heredity and Health in Africa (H3Africa) Consortium.” H3Africa Guideline for Informed Consent. 2013

